# Protocol and statistical analysis plan for the Mode of Ventilation During Critical IllnEss (MODE) trial

**DOI:** 10.1101/2023.07.21.23292998

**Authors:** Kevin P. Seitz, Bradley D. Lloyd, Li Wang, Matthew S. Shotwell, Edward T. Qian, Roger K. Richardson, Jeffery C. Rooks, Vanessa Hennings-Williams, Claire E. Sandoval, Whitney D. Richardson, Tracy Morgan, Amber N. Thompson, Pamela G. Hastings, Terry P. Ring, Joanna L. Stollings, Erica M. Talbot, David J. Krasinski, Bailey Decoursey, Kevin W. Gibbs, Wesley H. Self, Amanda S. Mixon, Todd W. Rice, Matthew W. Semler, Jonathan D. Casey

**Author notes:** **Corresponding author:** Kevin P. Seitz, MD, MSc, Address: 1121 21^1920^ Ave S, #T-1218, Nashville, TN, 37212. Authors contributed equally (Matthew W. Semler, Jonathan D. Casey). **Author contributions:** All authors approved the final version of this manuscript. Study concept and design: K.P.S., B.D.L., L.W., E.T.Q., R.K.R., J.C.R., W.H.S., T.W.R., M.W.S., J.D.C.; Acquisition of data: K.P.S., B.D.L., L.W., E.T.Q., C.E.S., W.D.R., T.M., A.N.T., P.G.H., T.P.R., J.L.S., E.M.T., D.J.K., B.D.; Drafting of the manuscript: K.P.S., M.W.S., J.D.C.; Critical revision of the manuscript for important intellectual content: All authors; Study supervision: K.P.S., A.S.M., T.W.R., M.W.S., J.D.C. **Conflicts of interest and financial disclosures:** J.D.C. reports a travel support from Fisher & Paykel Healthcare. T.W.R. reports unrelated consultant relationships with Cumberland Pharmaceuticals, Inc as Director of Medical Affairs, Cytovale, Inc, and Sanofi, Inc as a member of a DSMB. Supplemental digital content is available for this article.

## Abstract

**Introduction:** For every critically ill adult receiving invasive mechanical ventilation, clinicians must select a mode of ventilation. The mode of ventilation determines whether the ventilator directly controls the tidal volume or the inspiratory pressure. Newer hybrid modes allow clinicians to set a target tidal volume, for which the ventilator controls and adjusts the inspiratory pressure. A strategy of low tidal volumes and low plateau pressure improves outcomes, but the optimal mode to achieve these targets is not known.

**Methods and analysis:** The Mode of Ventilation During Critical Illness (MODE) trial is a cluster-randomized, multiple-crossover pilot trial being conducted in the medical intensive care unit (ICU) at an academic center. The MODE trial compares the use of volume control, pressure control, and adaptive pressure control. The study ICU is assigned to a single ventilator mode (volume control versus pressure control versus adaptive pressure control) for continuous mandatory ventilation during each 1-month study block. The assigned mode switches every month in a randomly generated sequence. The primary outcome is ventilator-free days (VFDs) to study day 28, defined as the number of days alive and free of invasive mechanical ventilation from the final receipt of mechanical ventilation to 28 days after enrollment. Enrollment began November 1, 2022 and will end on July 31, 2023.

**Ethics and dissemination:** The trial was approved by the Vanderbilt University Medical Center institutional review board (IRB# 220446). Results of this study will be submitted to a peer-reviewed journal and presented at scientific conferences.

**Trial registration number:** The trial was registered with clinicaltrials.gov on October 3, 2022, prior to initiation of patient enrollment on November 1, 2022 (ClinicalTrials.gov identifier: NCT05563779)

## Introduction

Approximately 2-3 million critically ill adults receive invasive mechanical ventilation in an intensive care unit (ICU) each year.^1–3^ While clinical trials have demonstrated that the use of smaller tidal volumes and lower plateau pressures can improve the outcomes of critically ill adults receiving invasive mechanical ventilation,^4, 5^ the optimal ventilator mode to achieve these targets remains unknown.

The tidal volume and the inspiratory airway pressure are directly related. For patients receiving invasive mechanical ventilation, delivering larger tidal volumes generates higher airway pressures and conversely, increasing airway pressures usually generates higher tidal volumes. Ventilator modes allow clinicians to control either the volume (“volume control” mode) or the pressure (“pressure control” mode) administered during inspiration.^6^ By controlling one variable, each mode provides indirect control over the other variable, which varies in a proportion dictated by the patient’s unique respiratory physiology. Modern ventilators also offer hybrid modes such as adaptive pressure control, a pressure targeted mode in which a clinician sets a target tidal volume (e.g., Pressure Regulated Volume Control) and the ventilator achieves that target by titrating the inspiratory pressure over multiple breaths to account for changes in patient effort and respiratory physiology.

Volume control, pressure control, and adaptive pressure control can all achieve low tidal volume and low plateau pressures, the key components of lung protective ventilation, a strategy proven to improve outcomes for critically ill adults receiving mechanical ventilation.^7–9^ Each mode differs, however, in the specifics of how it delivers inspiratory pressure, volume, and flow. These differences could theoretically affect pulmonary physiology, patient comfort, sedation requirements, risk of ventilator-induced lung injury, and clinical outcomes.^10, 11^ To date, no studies have found conclusive benefit from any mandatory mode of ventilation.^12^ Further, the only clinical trials comparing modes of mechanical ventilation among critically ill adults have been small and occurred when deep sedation and paralysis were common and spontaneous awakening and breathing trials had not yet become routine practice.^13, 14^

Given the absence of evidence regarding the ideal mode of mechanical ventilation, significant variation in the choice of mode exists in current clinical practice. Observational cohort studies in the United States and internationally have demonstrated that volume control, pressure control, and adaptive pressure control are each used routinely in the management of critically ill adults.^7, 8, 15^ Because the mode of mechanical ventilation must be set for every critically ill patient receiving mechanical ventilation and the relationship between the commonly used modes and patient outcomes remains uncertain, a large, randomized trial at multiple centers is needed to inform the optimal mode of mechanical ventilation for critically ill adults.^16–18^ Before such a trial can be conducted, additional data are needed regarding the feasibility of assigning the mode of mechanical ventilation for critically ill adults, of maintaining adherence to group assignment, and of using days alive and free of ventilation as a primary outcome for a trial comparing ventilator modes.^19–22^ To address this lack of preliminary data, we designed the Mode of Ventilation During Critical Illness (MODE) trial as a prospective, randomized, pilot trial comparing modes of continuous mandatory ventilation among critically ill adults in a medical ICU.

## METHODS and ANALYSIS

This manuscript was written by the MODE trial investigators, in accordance with Standard Protocol Items: Recommendations for Interventional Trials (SPIRIT) guidelines (online supplement, section 1). In this manuscript, we describe key elements of the trial protocol and statistical analysis plan.

The online supplementary materials provide additional background on: design decisions (section 2, 19, 22), mode monitoring and management (sections 3, 4, 12, 15, 23, 24), institutional protocols and management of critically ill adults receiving invasive mechanical ventilation (sections 5-11), a complete list of data elements (section 13), additional outcome definitions (section 14), details of the interim analysis (section 16), and secondary analysis considerations (section 17, 18).

### Study design

The MODE trial is a prospective, cluster-randomized, multiple-crossover trial being conducted in the medical ICU at a single center. This trial compares use of volume control versus pressure control versus adaptive pressure control among patients receiving invasive mechanical ventilation in the study ICU. Consistent with the goals of a pragmatic trial, delivery of the intervention is embedded in routine clinical care and managed by bedside clinicians. The primary outcome is the number of days alive and free of invasive mechanical ventilation to study day 28 after enrollment.

Feasibility measures are reported to inform the conduct of a subsequent multi-center trial. The trial protocol was approved by the Vanderbilt University Medical Center Institutional Review Board (IRB 220446) on July 21, 2022 and was registered with ClinicalTrials.gov (NCT05563779) on October 3, 2022, prior to initiation of patient enrollment on November 1, 2022.

### Study site and population

The trial is being conducted in the medical ICU at a tertiary-care teaching hospital. Patients are enrolled in the trial or excluded from the trial at the time of first receipt of invasive mechanical ventilation in the study ICU using the following criteria (Table 2):

### The inclusion criteria are

1. Age ≥ 18 years
2. Receiving mechanical ventilation through an endotracheal tube or tracheostomy
3. Admitted to the study ICU *The exclusion criteria are:*
4. Patient is pregnant
5. Patient is a prisoner
6. Patient receiving invasive mechanical ventilation at place of residence prior to hospital admission
7. Patient receiving extracorporeal membrane oxygenation at the time of admission to the study ICU.

### Randomization and treatment allocation

During each one-month period of the 9 months of enrollment in the MODE trial, the study ICU is assigned to use one of the three modes of mechanical ventilation (volume control vs pressure control vs adaptive pressure control). Each month, the ICU will switch between the three modes in a randomly generated sequence. The order of the study group assignments was generated by computerized randomization using permuted blocks of 3 to minimize the impact of seasonal variation. Patients will be analyzed in the group to which they were assigned at enrollment (intention-to-treat) even if they remain in the study ICU during a transition from one month to the next (“crossover”).

### Washout periods

The last 3 days of each month are an analytic washout period during which the study ICU continues to use the assigned ventilator mode, but new patients are not included in the primary analysis. The 3-day washout period will reduce the number of patients who experience a “crossover” from one assigned mode to another assigned mode in the 72-hour time window when feasibility outcomes of adherence are assessed.

### Study Interventions

#### Ventilator Mode

The MODE trial compares volume control, pressure control, and adaptive pressure control (Table 1). On initiation of invasive mechanical ventilation, nearly all patients require a ventilator mode that initiates breaths at a set frequency (mandatory mode of ventilation). The three most commonly used modes of mandatory ventilation are volume control, pressure control, and adaptive pressure control. Each of these modes provides continuous mandatory ventilation, in which every inspiratory effort by a patient triggers a machine-cycled breath delivered by the ventilator, and a set minimum respiratory rate is maintained by machine-triggered breaths as needed.^6^

**Table 1.** Characteristics of the modes of mechanical ventilation.

| Group Assignment | Volume Control | Pressure Control | Adaptive Pressure Control |
| --- | --- | --- | --- |
| Ventilator Mode Designation | *Volume Control* | *Pressure Control* | *Pressure Regulated Volume Control* |
| Control Variable | Volume | Pressure | Pressure |
| Breath Sequence | Continuous mandatory ventilation | Continuous mandatory ventilation | Continuous mandatory ventilation |
| Targeting Scheme | Set-point<br>(clinician sets target volume) | Set-point<br>(clinician sets target inspiratory pressure) | Adaptive<br>(clinician sets target volume and ventilator titrates pressure to achieve target volume) |

**Table 2.**
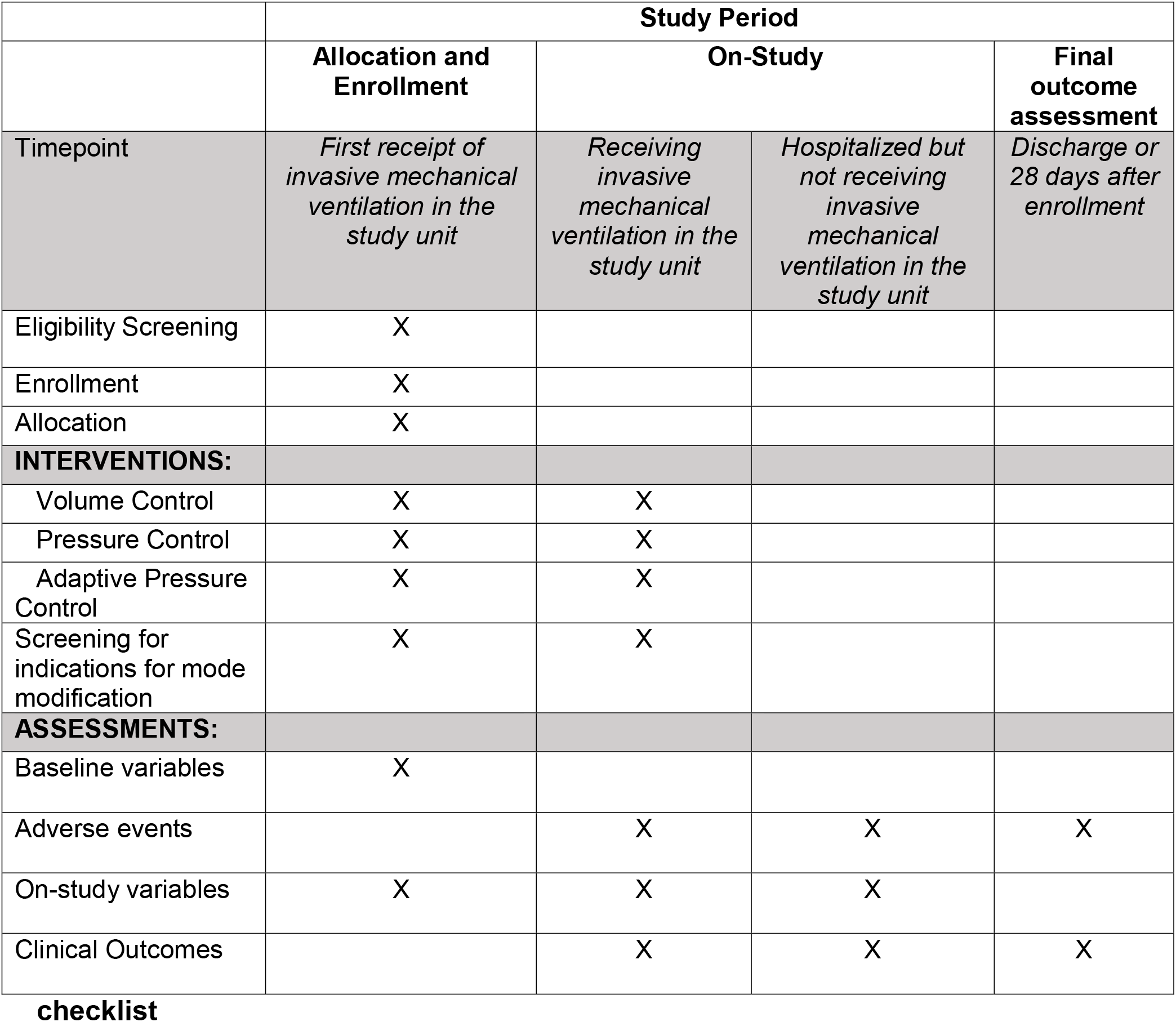
Standard Protocol Items: Recommendations for Interventional Trials.

|  | Study Period |  |  |  |
| --- | --- | --- | --- | --- |
|  | Allocation and Enrollment | On-Study |  | Final outcome assessment |
| Timepoint | *First receipt of invasive mechanical ventilation in the study unit* | *Receiving invasive mechanical ventilation in the study unit* | *Hospitalized but not receiving invasive mechanical ventilation in the study unit* | *Discharge or 28 days after enrollment* |
| Eligibility Screening | X |  |  |  |
| Enrollment | X |  |  |  |
| Allocation | X |  |  |  |
| **INTERVENTIONS:** |  |  |  |  |
| Volume Control | X | X |  |  |
| Pressure Control | X | X |  |  |
| Adaptive Pressure Control | X | X |  |  |
| Screening for indications for mode modification | X | X |  |  |
| **ASSESSMENTS:** |  |  |  |  |
| Baseline variables | X |  |  |  |
| Adverse events |  | X | X | X |
| On-study variables | X | X | X |  |
| Clinical Outcomes |  | X | X | X |

For patients enrolled in the MODE trial, clinicians are instructed to use the assigned mode beginning at the first receipt of invasive mechanical ventilation in the study ICU and ending at the first of: (1) extubation from mechanical ventilation, (2) transfer out of the study ICU, (3) treating clinician deciding that optimal care requires a different mode (with completion of a ventilator mode modification sheet), or (4) end of the one-month study block (online supplement, section 15). Any patients who remain mechanically ventilated during a “crossover” from one month to the next will, after the “crossover”, have their mode of ventilation determined by the clinical team. If a patient is enrolled, extubated, and re-intubated during the same study block, the study protocol will determine the ventilator mode until they meet one of the criteria listed above. The study protocol does not determine the ventilator mode during time-periods in which the patient is not receiving a mandatory mode of ventilation (e.g., use of pressure support ventilation during a spontaneous breathing trial), is not physically located in the study ICU (e.g., during transport), or is undergoing an invasive procedure (e.g., bronchoscopy).

In the study ICU, respiratory therapists typically have primary responsibility for determining the initial settings for invasive mechanical ventilation and titrating the settings to achieve clinical goals (e.g., SaO_2_, pCO_2_, target tidal volume, target plateau pressure, etc.). To set and titrate mechanical ventilators, respiratory therapists use standard-of-care clinical protocols jointly developed by respiratory therapy and physician leaders. For patients enrolled in the MODE trial, respiratory therapists employ preexisting clinical protocols dictating how each study group (volume control, pressure control, and adaptive pressure control) should be set and titrated (online supplement, section 5).

#### Ventilator Mode Modification Sheet

If at any time, any member of the clinical team, such as a respiratory therapist, advanced practice provider, or physician, determines a specific mandatory mode of ventilation is needed for the optimal treatment of any specific patient, they may use that mode of mechanical ventilation. In these cases, a one-page mode modification sheet is completed documenting the date, time, reason for modification, and the mode of ventilation chosen. Anticipated examples of conditions for which treating clinicians may elect to override the assigned mode include: refractory hypoxemia, persistently high peak pressures (> 40 cm H_2_O), dyssynchrony not amenable to changes within the assigned mode, excessive work of breathing, barotrauma, inability to limit tidal volumes delivered, or intrinsic positive end-expiratory pressure (PEEP). Strategies used to monitor and improve adherence to the assigned study mode are included in the online supplement (section 3, 4)

#### Co-interventions

For all patients, regardless of group assignment, existing ventilator protocols in the study ICU will be used to target a tidal volume of 6 mL/kg predicted body weight and a plateau pressure less than 30 cm H_2_O, while maintaining a respiratory rate less than 35 and pH of greater than 7.15, with a target of 7.30 to 7.45.^5^

On each day of mechanical ventilation, all patients in the study ICU are assessed for safety of a spontaneous awakening trial (SAT) and spontaneous breathing trial (SBT), and if safe, these procedures are performed, per the existing unit clinical protocols. As such, the SAT and SBT procedures are handled in the same way for each patient, regardless of study group assignment. For patients who have passed an SAT and SBT, the decision to discontinue invasive mechanical ventilation is made by the treating clinicians. Additional details regarding clinical practice guidelines in use are provided in the supplement (sections 6-12)

### Blinding

Consistent with prior trials evaluating mechanical ventilation settings,^5, 23^ patients and clinicians in the MODE trial are not blinded to trial group assignment.

### Data Collection

Trial personnel collect data by two methods to minimize observer bias. First, trial personnel review the electronic health record at least twice daily to confirm eligibility criteria and enrollment status for new patients, monitor adherence to the assigned ventilator mode, and screen for the occurrence of adverse events (online supplement, section 3). Trial personnel manually collect data on baseline characteristics, on study-management, and clinical outcomes.

Second, structured data recorded in routine clinical care is exported from the institution’s electronic health record into an Enterprise Data Warehouse. This method of data collection has been validated and used in prior pragmatic trials at this site.^23, 24^

The primary outcome is collected both by manual chart review and automated data collection for confirmation. Discordance between manual and automated data will be reviewed and manually adjudicated by a second investigator. The list of all variables collected is available in the online supplement (section 13).

### Outcomes

#### Primary Outcome

The primary outcome is the number of ventilator free days (VFDs) through day 28 after enrollment. VFDs will be defined as the number of whole calendar days alive and free of invasive mechanical ventilation from the final receipt of invasive mechanical ventilation through day 28 after enrollment.^25, 26^ Patients who die prior to hospital discharge on or before day 28 will receive 0 VFDs. Patients whose final receipt of invasive mechanical ventilation occurs on the day of enrollment (day 1) and survive to day 28 will receive 27 VFDs. Additional detail is provided in the online supplement (section 20).

#### Additional Outcomes

Feasibility outcomes, exploratory efficacy and safety outcomes, and clinical outcomes were pre-specified and are described in Table 3.

**Table 3.** Feasibility and Exploratory Outcomes.

|  |  |
| --- | --- |
| **Feasibility Outcomes**<br>*Prespecified feasibility outcomes focus on adherence to group assignment and separation between groups in mode of mechanical ventilation.* |  |
| 1 | Exposure to assigned study mode in the first 3 days: proportion of time in the assigned mode while receiving invasive mechanical ventilation in the study ICU between enrollment and 72 hours after enrollment |
| 2 | Adherence to study mode in first 3 days: proportion of time in the assigned mode while receiving invasive mechanical ventilation in the study ICU with a mandatory mode between enrollment and 72 hours after enrollment (excluding time spent in spontaneous modes) |
| 3 | Time from enrollment to initiation of assigned mode of mechanical ventilation |
| 4 | Receipt of a “Mode Modification Sheet” completed by treating clinicians |
| **Exploratory Efficacy and Safety Outcomes**<br>*Exploratory outcomes for efficacy and safety were prespecified to provide intermediate measures that may be affected by the intervention (ventilator mode) and may affect the primary outcome (ventilator-free days to study day 28) but in this study are not meaningful in isolation. Additional outcomes are listed below, with definitions available in the online supplement (section 14).* |  |
| 1 | Median exhaled tidal volume (mL/kg predicted body weight) on each study day |
| 2 | Exhaled tidal volumes above target range: proportion of recorded breaths with exhaled tidal volume values above the target range (>8mL/kg predicted body weight) on each study day |
| 3 | Hypoxemia during mechanical ventilation: episodes of hypoxemia during mechanical ventilation: SpO<sub>2</sub> < 85% for more than 5 minutes |
| 4 | Severe acidemia during mechanical ventilation: episodes of severe acidemia during mechanical ventilation: pH < 7.1 on blood gas |
| 5 | Number of blood gas laboratory tests per day while receiving mechanical ventilation |
| 6 | Pneumomediastinum or pneumothorax during course of mechanical ventilation |
| 7 | SOFA score daily on the first 7 study days |
| 8 | Delirium and coma-free days to day 28 |
| **Exploratory Clinical Outcomes**<br>*More detailed definitions are available in the online supplement (section 14).* |  |
| 1 | ICU-free days to study day 28 |
| 2 | Hospital-free days to study day 28 |
| 3 | In-hospital mortality to study day 28 |

### Statistical Analysis and Reporting

#### Sample size estimation and power calculation

The objectives of this trial are to demonstrate the feasibility of comparing modes of invasive mechanical ventilation in critically ill adults using a cluster-randomized, multiple-crossover design and to collect preliminary data to inform a subsequent multi-center trial using a similar study design. To assess whether adherence to trial group assignment can be maintained across “crossovers” in the assigned mode of mechanical ventilation, we will need to observe two end-of-period “crossovers” into each mode while maintaining an equal number of treatment blocks for each mode. Doing so will require a total of 9 months. Based on data from a prior trial in the same ICU,^27^ we anticipate an average of 75 patients will be enrolled per month. In 9 months, we expect to enroll an estimated 675 patients, of whom 69 will be enrolled during washout periods and excluded from the primary analysis. As a result, approximately 606 patients will be included in the primary analysis.

While the sample size was determined to demonstrate feasibility of the cluster-crossover design, we anticipate this pilot will be the largest randomized trial of ventilator modes to date, and as such, we plan to compare ventilator free days to day 28 (primary outcome) among groups. In a prior cluster-randomized cluster-crossover trial in the same ICU,^27^ mechanically ventilated patients experienced a median of 22 VFDs [IQR 0-25 VFDs] and an intracluster, intraperiod correlation of 0.01. With 606 patients (202 in each group) in the primary analysis, a standard deviation in the primary outcome of VFDs of 11 days, and a two-sided alpha of 0.05, the MODE trial will have 80 percent statistical power to detect an absolute difference between groups in the primary outcome of 3 ventilator-free days.

### Data and Safety Monitoring Board (DSMB) and interim analysis

An independent Data Safety and Monitoring Board (DSMB) oversees conduct of the MODE trial. The DSMB is composed of three physicians with expertise in critical care medicine, pulmonary medicine, biostatistics, and clinical trials. On March 22, 2023, the DSMB conducted a single, planned interim analysis for safety after the first 3 months of enrollment. The DSMB recommended that the trial continue without modification. Details of the safety analysis are provided in the online supplement (section 16). Given that the goal of the study is to demonstrate feasibility rather than demonstrating efficacy, there was no early stopping criterion for efficacy or futility.

### Statistical analysis principles

Statistical analyses will be completed using R (R foundation for Statistical Computing, Vienna, Austria), and analyses will be done at the level of an individual patient during an individual hospitalization in an intention-to-treat fashion.

### Main analysis of the primary outcome

The sole pre-specified primary outcome of ventilator free days will be compared among the three trial groups in an intention-to-treat fashion among all patients enrolled in the trial except those enrolled during one of the 3-day washout periods. We will compare the primary outcome of ventilator-free days between the trial groups using a proportional odds model with independent variables of group assignment (volume control, pressure control, or adaptive pressure control) and time. Time (in days) will be treated as a continuous variable with values ranging from 1 (first day of enrollment) to 272 (final day of enrollment) and will be analyzed using restricted cubic splines with multiple knots to allow for non-linearity resulting from seasonality or secular trends. A p-value threshold of 0.05 will be considered significant evidence of an overall difference across treatment groups. In addition to assessing for overall differences across the three groups, we will estimate the differences between each pair of modes by extracting odds ratios with 95% confidence intervals from the model. Sensitivity analyses of the primary outcome are described in the online supplement (section 18).

### Analysis of effect modification for the primary outcome

We will examine whether pre-specified baseline variables modify the effect of study group on the primary outcome using tests of statistical interaction in a proportional odds model. Independent variables will include study group assignment, the potential effect modifier of interest, the interaction between the two (e.g., study group * shock), and time. Continuous variables will be analyzed using restricted cubic splines to allow for non-linear relationships. Significance will be determined by the P value for the interaction term(s), with values less than 0.05 considered significant evidence of an interaction.

In accordance with the Instrument for assessing the Credibility of effect Modification Analyses (ICEMAN) recommendations, we prespecified the following baseline variables as potential modifiers of the effect of study group on the primary outcome and hypothesized the direction of the effect modification for each in the supplement (section 17):

1. Age (continuous variable)
2. Duration of invasive mechanical ventilation prior to enrollment (0 minutes; 1 to 360 minutes; >360 minutes)
3. Pre-enrollment fraction of inspired oxygen (FiO_2_) (continuous variable).
4. SOFA score at enrollment (continuous variable)
5. Shock receiving vasopressors (yes, no)
6. Indications for intubation (categories are not mutually exclusive)

a. Hypoxemic respiratory failure (yes, no)
b. Hypercarbic respiratory failure (yes, no)
c. Altered mental status or airway protection (yes, no)
7. Chronic Obstructive Pulmonary Disease (yes, no)

### Analysis of the exploratory outcomes

Each of the exploratory outcomes will be compared between groups in an intention-to-treat fashion with an approach similar to that used for the primary outcome. A logistic model will be used for binary outcomes, and a proportional odds model for ordinal and continuous outcomes. All models will include independent covariates of group assignment and time.

### Trial status

The MODE trial is an ongoing trial comparing volume control versus pressure control versus adaptive pressure control for critically ill adults receiving invasive mechanical ventilation. Patient enrollment began on November 1, 2022 and is anticipated to conclude on July 31, 2023.

### ETHICS and DISSEMINATION

#### Waiver of Informed Consent

Volume control, pressure control, and adaptive pressure control are all common approaches to controlled mechanical ventilation for critically ill adults. All represent standard-of-care treatments in current clinical practice. Because the study involves minimal incremental risk and obtaining informed consent would be impracticable, this trial was approved by the Vanderbilt University Medical Center IRB with a waiver of informed consent (#220446). Additional rationale is provided in the online supplement (section 20).

### Protocol changes

All changes to the trial protocol will be distributed to the IRB and recorded on ClinicalTrials.gov as per SPIRIT guidelines (online supplement, section 21).

### Data handling

Privacy protocols and data handling are reported in the online supplement (section 22).

### Dissemination plan

Investigators will submit the MODE trial results to a peer-reviewed journal for consideration of publication, and results will be presented at scientific conferences.

## CONCLUSION

The MODE trial will provide the best evidence to date regarding the effect of mode of mechanical ventilation on the clinical outcomes of critically ill adults and will provide data regarding the feasibility for a definitive multi-center trial. To aid in the transparency and interpretation of trial results, this protocol and statistical analysis plan has been finalized prior to the conclusion of patient enrollment.

## Data Availability

At the time of publication of the trial described in this protocol, a de-identified version of the database will be generated and available upon reasonable request to the authors.

---

**Figure 1.**
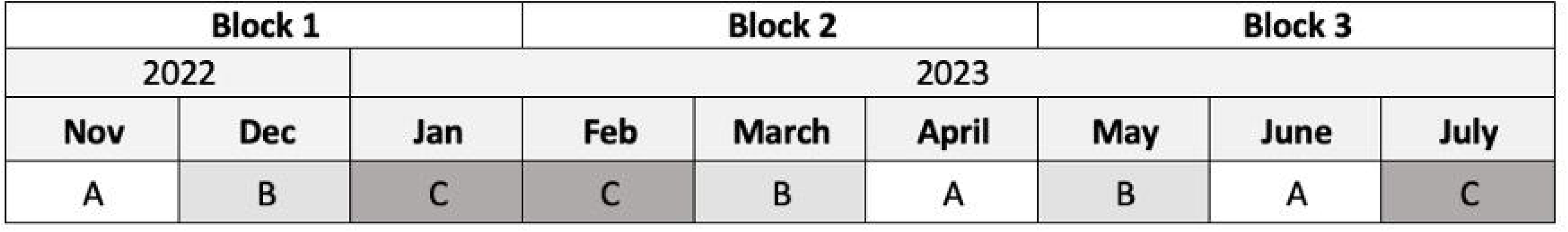
Group assignment during the MODE trial. For each of the 1-month study periods, the study unit is randomly assigned to a study mode for continuous mandatory ventilation. The letters ‘A’, ‘B’, and ‘C’ each correspond to one of the three study modes.

## REFERENCES

1. Wunsch H, Angus DC, Harrison DA, Linde-Zwirble WT, Rowan KM. Comparison of Medical Admissions to Intensive Care Units in the United States and United Kingdom. Am J Respir Crit Care Med 2011;183(12):1666–73.

2. Wunsch H, Linde-Zwirble WT, Angus DC, Hartman ME, Milbrandt EB, Kahn JM. The epidemiology of mechanical ventilation use in the United States*: Critical Care Medicine 2010;38(10):1947–53.

3. Adhikari NK, Fowler RA, Bhagwanjee S, Rubenfeld GD. Critical care and the global burden of critical illness in adults. The Lancet 2010;376(9749):1339–46.

4. Writing Group for the PReVENT Investigators, Simonis FD, Serpa Neto A, et al. Effect of a Low vs Intermediate Tidal Volume Strategy on Ventilator-Free Days in Intensive Care Unit Patients Without ARDS: A Randomized Clinical Trial. JAMA 2018;320(18):1872.

5. Acute Respiratory Distress Syndrome Network. Ventilation with Lower Tidal Volumes as Compared with Traditional Tidal Volumes for Acute Lung Injury and the Acute Respiratory Distress Syndrome. N Engl J Med 2000;342(18):1301–8.

6. Chatburn RL, El-Khatib M, Mireles-Cabodevila E. A Taxonomy for Mechanical Ventilation: 10 Fundamental Maxims. Respiratory Care 2014;59(11):1747–63.

7. Qadir N, Bartz RR, Cooter ML, et al. Variation in Early Management Practices in Moderate-to-Severe ARDS in the United States. Chest 2021;S0012369221010783.

8. Neto AS, Barbas CSV, Simonis FD, et al. Epidemiological characteristics, practice of ventilation, and clinical outcome in patients at risk of acute respiratory distress syndrome in intensive care units from 16 countries (PRoVENT): an international, multicentre, prospective study. The Lancet Respiratory Medicine 2016;4(11):882– 93.

9. Meade MO, Cook DJ, Guyatt GH, et al. Ventilation strategy using low tidal volumes, recruitment maneuvers, and high positive end-expiratory pressure for acute lung injury and acute respiratory distress syndrome: a randomized controlled trial. JAMA 2008;299(6):637–45.

10. Gibbs KW, Forbes JL, O’Connell NS, et al. Excess Tidal Volume Ventilation in Critically Ill Adults Receiving Adaptive Pressure Control: A Cohort Study. Annals ATS 2022;AnnalsATS.202203-200RL.

11. Beitler JR, Sands SA, Loring SH, et al. Quantifying unintended exposure to high tidal volumes from breath stacking dyssynchrony in ARDS: the BREATHE Criteria. Intensive Care Med 2016;42(9):1427–36.

12. Rittayamai N, Katsios CM, Beloncle F, Friedrich JO, Mancebo J, Brochard L. Pressure-Controlled vs Volume-Controlled Ventilation in Acute Respiratory Failure. Chest 2015;148(2):340–55.

13. Esteban A, Alía I, Gordo F, et al. Prospective Randomized Trial Comparing Pressure-Controlled Ventilation and Volume-Controlled Ventilation in ARDS. Chest 2000;117(6):1690–6.

14. Rappaport SH, Shpiner R, Yoshihara G, Wright J, Chang P, Abraham E. Randomized, prospective trial of pressure-limited versus volume-controlled ventilation in severe respiratory failure. Crit Care Med 1994;22(1):22–32.

15. Lanspa MJ, Gong MN, Schoenfeld DA, et al. Prospective Assessment of the Feasibility of a Trial of Low Tidal Volume Ventilation for Patients with Acute Respiratory Failure. Annals ATS 2018;AnnalsATS.201807-459OC.

16. Marini JJ. Point: Is Pressure Assist-Control Preferred Over Volume Assist-Control Mode for Lung Protective Ventilation in Patients With ARDS? Yes. Chest 2011;140(2):286–90.

17. MacIntyre N. Counterpoint: Is Pressure Assist-Control Preferred Over Volume Assist-Control Mode for Lung Protective Ventilation in Patients With ARDS? No. Chest 2011;140(2):290–2.

18. Branson R. Should Adaptive Pressure Control Modes Be Utilized for Virtually All Patients Receiving Mechanical Ventilation? RESPIRATORY CARE 2007;52(4):11.

19. Moore CG, Carter RE, Nietert PJ, Stewart PW. Recommendations for Planning Pilot Studies in Clinical and Translational Research. Clin Transl Sci 2011;4(5):332– 7.

20. Taylor SP, Kowalkowski MA. Using Implementation Science-Guided Pilot Studies to Assess and Improve the Informativeness of Clinical Trials. J GEN INTERN MED 2021;36(2):533–6.

21. Eldridge SM, Chan CL, Campbell MJ, et al. CONSORT 2010 statement: extension to randomised pilot and feasibility trials. BMJ 2016;i5239.

22. Gibbs KW, Forbes JL, Harrison KJ, et al. A Pragmatic Pilot Trial Comparing Patient-Triggered Adaptive Pressure Control to Patient-Triggered Volume Control Ventilation in Critically Ill Adults. Respir Care 2023;respcare.10803.

23. Semler MW, Casey JD, Lloyd BD, et al. Oxygen-Saturation Targets for Critically Ill Adults Receiving Mechanical Ventilation. N Engl J Med 2022;NEJMoa2208415.

24. Semler MW, Rice TW, Shaw AD, et al. Identification of Major Adverse Kidney Events Within the Electronic Health Record. J Med Syst 2016;40(7):167.

25. Schoenfeld DA, Bernard GR. Statistical evaluation of ventilator-free days as an efficacy measure in clinical trials of treatments for acute respiratory distress syndrome: Critical Care Medicine 2002;30(8):1772–7.

26. Harhay MO, Wagner J, Ratcliffe SJ, et al. Outcomes and Statistical Power in Adult Critical Care Randomized Trials. Am J Respir Crit Care Med 2014;189(12):1469– 78.

27. Semler MW, Self WH, Wanderer JP, et al. Balanced Crystalloids versus Saline in Critically Ill Adults. N Engl J Med 2018;378(9):829–39.

