## Supplementary material for "Protocol and statistical analysis plan for the Mode of Ventilation During Critical IllnEss (MODE) trial": Online Supplemental Materials

### SUPPLEMENTAL METHODS

### 1. Spirit 2013 Checklist

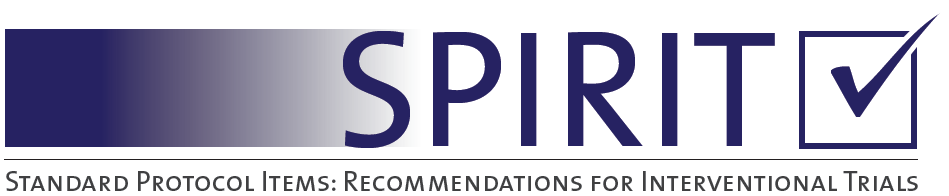

SPIRIT 2013 Checklist: Recommended items to address in a clinical trial protocol and related documents*

| Section/item | Item No | Description | Addressed on page number |
| --- | --- | --- | --- |
| **Administrative information** | | |  |
| Title | 1 | Descriptive title identifying the study design, population, interventions, and, if applicable, trial acronym | 1 |
| Trial registration | 2a | Trial identifier and registry name. If not yet registered, name of intended registry | 6,10 |
|  | 2b | All items from the World Health Organization Trial Registration Data Set | 1-3, 8,10-12, 15-16, 18, 24-25 |
| Protocol version | 3 | Date and version identifier | 1 |
| Funding | 4 | Sources and types of financial, material, and other support | 2-3 |
| Roles and responsibilities | 5a | Names, affiliations, and roles of protocol contributors | 2 |
|  | 5b | Name and contact information for the trial sponsor | NA |
|  | 5c | Role of study sponsor and funders, if any, in study design; collection, management, analysis, and interpretation of data; writing of the report; and the decision to submit the report for publication, including whether they will have ultimate authority over any of these activities | NA |
|  | 5d | Composition, roles, and responsibilities of the coordinating centre, steering committee, endpoint adjudication committee, data management team, and other individuals or groups overseeing the trial, if applicable (see Item 21a for data monitoring committee) | 2 |
| Introduction |  |  |  |
| Background and rationale | 6a | Description of research question and justification for undertaking the trial, including summary of relevant studies (published and unpublished) examining benefits and harms for each intervention | 7-8 |
|  | 6b | Explanation for choice of comparators | 11-12 |
| Objectives | 7 | Specific objectives or hypotheses | 8 |
| Trial design | 8 | Description of trial design including type of trial (eg, parallel group, crossover, factorial, single group), allocation ratio, and framework (eg, superiority, equivalence, noninferiority, exploratory) | 9 |
| Methods: Participants, interventions, and outcomes | | |  |
| Study setting | 9 | Description of study settings (eg, community clinic, academic hospital) and list of countries where data will be collected. Reference to where list of study sites can be obtained | 10 |
| Eligibility criteria | 10 | Inclusion and exclusion criteria for participants. If applicable, eligibility criteria for study centres and individuals who will perform the interventions (eg, surgeons, psychotherapists) | 10 |
| Interventions | 11a | Interventions for each group with sufficient detail to allow replication, including how and when they will be administered | 11-14 |
|  | 11b | Criteria for discontinuing or modifying allocated interventions for a given trial participant (eg, drug dose change in response to harms, participant request, or improving/worsening disease) | 13 |
|  | 11c | Strategies to improve adherence to intervention protocols, and any procedures for monitoring adherence (eg, drug tablet return, laboratory tests) | 14, Sup 33 |
|  | 11d | Relevant concomitant care and interventions that are permitted or prohibited during the trial | 13-14 |
| Outcomes | 12 | Primary, secondary, and other outcomes, including the specific measurement variable (eg, systolic blood pressure), analysis metric (eg, change from baseline, final value, time to event), method of aggregation (eg, median, proportion), and time point for each outcome. Explanation of the clinical relevance of chosen efficacy and harm outcomes is strongly recommended | 15 |
| Participant timeline | 13 | Time schedule of enrolment, interventions (including any run-ins and washouts), assessments, and visits for participants. A schematic diagram is highly recommended (see Figure) | Figure 1 |
| Sample size | 14 | Estimated number of participants needed to achieve study objectives and how it was determined, including clinical and statistical assumptions supporting any sample size calculations | 16 |
| Recruitment | 15 | Strategies for achieving adequate participant enrolment to reach target sample size | 16 |
| **Methods: Assignment of interventions (for controlled trials)** | | |  |
| Allocation: |  |  |  |
| Sequence generation | 16a | Method of generating the allocation sequence (eg, computer-generated random numbers), and list of any factors for stratification. To reduce predictability of a random sequence, details of any planned restriction (eg, blocking) should be provided in a separate document that is unavailable to those who enrol participants or assign interventions | 10-11 |
| Allocation concealment mechanism | 16b | Mechanism of implementing the allocation sequence (eg, central telephone; sequentially numbered, opaque, sealed envelopes), describing any steps to conceal the sequence until interventions are assigned | 10-11, Figure 2 |
| Implementation | 16c | Who will generate the allocation sequence, who will enrol participants, and who will assign participants to interventions | 10-13 |
| Blinding (masking) | 17a | Who will be blinded after assignment to interventions (eg, trial participants, care providers, outcome assessors, data analysts), and how | 14 |
|  | 17b | If blinded, circumstances under which unblinding is permissible, and procedure for revealing a participant’s allocated intervention during the trial | NA |
| **Methods: Data collection, management, and analysis** | | |  |
| Data collection methods | 18a | Plans for assessment and collection of outcome, baseline, and other trial data, including any related processes to promote data quality (eg, duplicate measurements, training of assessors) and a description of study instruments (eg, questionnaires, laboratory tests) along with their reliability and validity, if known. Reference to where data collection forms can be found, if not in the protocol | 14-15 |
|  | 18b | Plans to promote participant retention and complete follow-up, including list of any outcome data to be collected for participants who discontinue or deviate from intervention protocols | 15 |
| Data management | 19 | Plans for data entry, coding, security, and storage, including any related processes to promote data quality (eg, double data entry; range checks for data values). Reference to where details of data management procedures can be found, if not in the protocol | 14-15, 20 |
| Statistical methods | 20a | Statistical methods for analysing primary and secondary outcomes. Reference to where other details of the statistical analysis plan can be found, if not in the protocol | 18-19 |
|  | 20b | Methods for any additional analyses (eg, subgroup and adjusted analyses) | 18-19 |
|  | 20c | Definition of analysis population relating to protocol non-adherence (eg, as randomised analysis), and any statistical methods to handle missing data (eg, multiple imputation) | 17-19, Sup 37-38 |
| **Methods: Monitoring** | | |  |
| Data monitoring | 21a | Composition of data monitoring committee (DMC); summary of its role and reporting structure; statement of whether it is independent from the sponsor and competing interests; and reference to where further details about its charter can be found, if not in the protocol. Alternatively, an explanation of why a DMC is not needed | 17 |
|  | 21b | Description of any interim analyses and stopping guidelines, including who will have access to these interim results and make the final decision to terminate the trial | 17 |
| Harms | 22 | Plans for collecting, assessing, reporting, and managing solicited and spontaneously reported adverse events and other unintended effects of trial interventions or trial conduct | 14, Sup 44-48 |
| Auditing | 23 | Frequency and procedures for auditing trial conduct, if any, and whether the process will be independent from investigators and the sponsor | Sup 44-48 |
| Ethics and dissemination | | |  |
| Research ethics approval | 24 | Plans for seeking research ethics committee/institutional review board (REC/IRB) approval | 20 |
| Protocol amendments | 25 | Plans for communicating important protocol modifications (eg, changes to eligibility criteria, outcomes, analyses) to relevant parties (eg, investigators, REC/IRBs, trial participants, trial registries, journals, regulators) | 20 |
| Consent or assent | 26a | Who will obtain informed consent or assent from potential trial participants or authorised surrogates, and how (see Item 32) | 20 |
|  | 26b | Additional consent provisions for collection and use of participant data and biological specimens in ancillary studies, if applicable | 20 |
| Confidentiality | 27 | How personal information about potential and enrolled participants will be collected, shared, and maintained in order to protect confidentiality before, during, and after the trial | Sup 43 |
| Declaration of interests | 28 | Financial and other competing interests for principal investigators for the overall trial and each study site | 2-3 |
| Access to data | 29 | Statement of who will have access to the final trial dataset, and disclosure of contractual agreements that limit such access for investigators | 20 |
| Ancillary and post-trial care | 30 | Provisions, if any, for ancillary and post-trial care, and for compensation to those who suffer harm from trial participation | NA |
| Dissemination policy | 31a | Plans for investigators and sponsor to communicate trial results to participants, healthcare professionals, the public, and other relevant groups (eg, via publication, reporting in results databases, or other data sharing arrangements), including any publication restrictions | 20 |
|  | 31b | Authorship eligibility guidelines and any intended use of professional writers | 1-2 |
|  | 31c | Plans, if any, for granting public access to the full protocol, participant-level dataset, and statistical code | Sup 43 |
| Appendices |  |  |  |
| Informed consent materials | 32 | Model consent form and other related documentation given to participants and authorised surrogates | NA |
| Biological specimens | 33 | Plans for collection, laboratory evaluation, and storage of biological specimens for genetic or molecular analysis in the current trial and for future use in ancillary studies, if applicable | NA |

*It is strongly recommended that this checklist be read in conjunction with the SPIRIT 2013 Explanation & Elaboration for important clarification on the items. Amendments to the protocol should be tracked and dated. The SPIRIT checklist is copyrighted by the SPIRIT Group under the Creative Commons “[Attribution-NonCommercial-NoDerivs 3.0 Unported](http://www.creativecommons.org/licenses/by-nc-nd/3.0/)” license.

### 2. Rationale for Cluster-Level Allocation

Group assignment in the MODE trial occurs at the level of the ICU (cluster) for multiple reasons. In routine clinical care in the study ICU, 2 to 4 respiratory therapists set the ventilator mode and adjust settings to maintain gas exchange and patient comfort for all mechanically ventilated adults, with input from nurses and physicians. The management of mechanical ventilation (selection of fraction of inspired oxygen, titration of positive end-expiratory pressure, screening for and performance of spontaneous breathing trials) is governed by unit-wide protocols implemented by the 2 to 4 respiratory therapists for all patients in the unit. Assigning the entire unit to a single ventilator mode both emulates the way mechanical ventilation is managed during clinical care and limits contamination that might result from a respiratory therapist managing multiple patients assigned to different modes of mechanical ventilation.

Additionally, mechanical ventilation can damage lungs even after brief periods of ventilation, and the initial settings have a unique significance. For example, during the minutes-to-hours of temporary mechanical ventilation for surgical procedures, harmful ventilator settings can affect organ function and clinical outcomes.^1^ In critically ill patients, the initial hours of invasive mechanical ventilation are also the period with the highest risk of worsening lung injury, dyssynchrony, increased work of breathing, and hemodynamic compromise. Prior research has demonstrated that [1] the very first ventilator settings at the time the patient is being initiated on invasive mechanical ventilation in the ICU are associated with differences in mortality^2,3^ and [2] the association between ventilator settings and mortality is larger for earlier settings compared to later settings.^4^ The ventilator protocol used to set the ventilator mode in the study ICU begins immediately at the time of initiation of mechanical ventilation. Enrollment immediately after initiation of invasive mechanical ventilation minimizes pre-study exposure to other modes and ventilator settings, and it facilitates on-study separation between groups.

Recognizing the importance of initial ventilator settings, three recent clinical trials have examined other ventilator settings (i.e. end-expiratory pressure, tidal volume, and SpO_2_ target) by attempting to enroll patients soon after the initiation of mechanical ventilation in the ICU. ^5–7^ In these patient-level, parallel-group trials, however, the logistical challenges of performing screening, enrollment, randomization, and study group assignment resulted in a significant gap between initiation of invasive mechanical ventilation and delivery of the respective trial interventions. Those approaches also led to the exclusion of 60-67% of eligible patients – raising concern for systematic exclusion of important patient groups (e.g., patients with higher acuity of illness). In MODE, group assignment at the cluster level allows enrollment immediately on initiation of invasive mechanical ventilation in the ICU. This approach emulates the manner in which ventilator settings are managed in practice, decreases pre-study exposure to harmful ventilator settings, facilitates early separation in the receipt of ventilator modes between groups, and precludes systematic exclusion of important patient groups.

### 3. Approach to Monitoring Ventilator Mode

For all mechanically ventilated patients in the study ICU, ventilators display settings such as mode of ventilation and measurements such as observed tidal volume on a digital display at the patient’s bedside in real time. For critical measurements such as respiratory rate, minute ventilation, and peak pressures, audible alarms from the ventilator alert clinicians to potentially dangerous situations.

These settings and measurements are frequently sampled and uploaded automatically into the electronic medical record frequently and are stored in the data warehouse. This allows for much more representative assessment of the proportion of time that patients receive each ventilator mode over the course of the day compared to prior trials which documented ventilator mode once or twice daily.^8,9^

### 4. Feedback on Study Mode Adherence

During the study, study personnel will monitor compliance with the assigned ventilator mode. Study personnel will remotely monitor ventilator modes up to four times daily from 5AM through 11PM during weekdays and twice daily during weekends to provide feedback to treating clinicians and explore reasons why patients are not receiving the assigned mode of mechanical ventilation. Study personnel will also physically visit the study ICU at least once daily during weekdays to solicit safety concerns and adverse events and to identify and address barriers to ventilator mode assignment compliance.

In addition, study personnel will attend respiratory therapy group meetings, nursing unit board meetings, and ICU physician meetings to educate staff about the study and receive input on conduct of the trial. This approach to daily monitoring and intermittent feedback to clinical personnel successfully achieved 80% compliance with the assigned intervention in a prior cluster-crossover trial of ventilator settings in the same ICU.10

### 5. Ventilator Management Guidelines in the Study ICU

SUMMARY OF VUMC MICU VENTILATOR PROTOCOL GUIDELINES:

**Ventilator Set Up and Adjustment:**

1. Calculate Predicted Body Weight (PBW):
   a. In males = 50 - 2.3 (height (inches) - 60)
   b. In females = 45.5 + 2.3 (height (inches) - 60)

2. Select Mode and initial default settings:

a. **Volume Control.** Set Tidal Volume (Vt) to target 6 mL/kg PBW. Set Flow Pattern: 100 or [square-wave icon]; Set T pause (s): 0.

b. **Pressure Regulated Volume Control.** Set Vt to target 6 mL/kg PBW.

c. **Pressure Control.** Set PC above PEEP to 15 cmH_2_O, set inspiratory time to 0.9 s and confirm delivered Vt is appropriate for target 6 mL/kg PBW.

3. Set initial respiratory rate (RR) to approximate baseline Minute Ventilation (12-16 bpm, not > 35 bpm).

4. Set alarms:

For **VC and PRVC**: High Peak Pressure alarm at 40 cm H_2_0

For **PC**: Low Minute Ventilation alarm at 2 L/min less than the current Minute Ventilation

5. Adjust Vt and RR to achieve pH and plateau pressure goals below.

**Lung Protective Ventilation Goals – Vt** ≤ 6 mL/kg PBW**, Pplat** < 30 cmH_2_0

**For Pressure Control, where Vt cannot be directly adjusted, adjust PC over PEEP in increments of 2 cmH_2_0 and monitor resulting Vt to achieve the changes outlined below.*

If Vt > 6 mL/kg PBW:

Reduce Vt by 1 mL/kg at intervals ≤ 2 hours until Vt = 6 mL/kg PBW.

If Pplat > 30 (or PIP > 35):

Decrease Vt by 1 mL/kg steps to a minimum of 4mL/kg PBW.

If Pplat < 25 and Vt < 6 mL/kg:

Increase Vt by 1 mL/kg until either Pplat > 25 or Vt = 6 mL/kg PBW.

If Pplat <3 0 and breath stacking occurs:

May increase Vt in 1 mL/kg increments to maximum of 8 mL/kg PBW.

**Ventilation Goals - pH**: 7.30-7.45

If pH 7.15-7.30:

Increase RR until pH > 7.30 or PaCO2 < 25 (Max RR = 35).

If RR = 35 and PaCO2 < 25:

May give NaHCO_3_, to be managed by primary team.

If pH < 7.15:

Increase RR to 35.

If pH remains < 7 .15 and NaHCO_3_ considered or infused:

Vt may be increased until pH >7.15 or Vt = 8 mL/kg PBW (Pplat target may be exceeded).

If pH >7.45:

Decrease RR rate.

**Oxygenation goals -** **SpO_2_:** 88-95% or **PaO_2_:** 55-80 mmHg.

Use incremental FiO_2_/PEEP combinations below to provide minimum FiO_2_ and PEEP necessary to achieve PaO_2_ or SpO_2_ goals.

| **FiO2** | 0.3-0.4 | 0.4 | 0.5 | 0.5 | 0.6 | 0.7 | 0.7 | 0.7 | 0.8 | 0.9 | 0.9 | 0.9 | 1 |
| --- | --- | --- | --- | --- | --- | --- | --- | --- | --- | --- | --- | --- | --- |
| **PEEP** | 5 | 8 | 8 | 10 | 10 | 10 | 12 | 14 | 14 | 14 | 16 | 18 | 18-25 |

**Other Settings**

**I:E ratio goal:** 1:1.0 – 1:3.0. Adjust Ti to achieve goal.

If FiO_2_ = 1.0 and PEEP = 24 cmH_2_0, may adjust I:E to 1:1.

Avoid inverse-ratio ventilation.

**Trigger:** 1.6 L/min by default, can be increased or reduced to improve synchrony

**Tinsp rise (s):** 0.15 s by default, can be increased or decreased to improve ventilation

**Monitoring and Documentation:**

Measure & record Pplat with inspiratory pause of at least 0.5 s, SpO_2_, Total RR, Vt and pH (if available) at least every 4 hours AND after each change in PEEP, Vt, or PC above PEEP settings.

Once the patient tolerates FiO_2_ < 0.5 and PEEP < 8 cmH_2_O, advance to Part II: Spontaneous Breathing Trial Readiness Assessment and Ventilator Discontinuation Guidelines.

**Table for Ventilator Management Guidelines in the Study ICU**

|  | **Volume Control** | **Pressure Control** | **Adaptive Pressure Control** |
| --- | --- | --- | --- |
| Mode option | Volume Control | Pressure Control | Pressure Regulated Volume Control (PRVC) |
| Tidal Volume target (mL/kg PBW) | 6 | 6 | 6 |
| Tidal Volume range (mL/kg PBW) | 4 - 8 | 4 - 8 | 4 - 8 |
| Initial breath settings | Tidal volume to 6 mL/kg PBW | PC over PEEP to 15 cm H2O, Insp Time to 0.9s | Tidal volume to 6 mL/kg PBW |
| Achieving target Tidal Volume | Directly adjust tidal volume | Dependent on adjustment of PC over PEEP | Directly adjust tidal volume target |
| Plat Pressure target (cm H20) | ≤ 30 | ≤ 30 | ≤ 30 |
| Achieving target Plateau Pressure | Dependent on adjustment of tidal volume | Directly adjust PC over PEEP | Dependent on adjustment of  tidal volume |
| Flow | Set directly | Dependent ventilator and patient factors | Dependent ventilator and patient factors |
| Flow Pattern | Square waveform | Decelerating, dynamic | Decelerating, dynamic |
| Arterial pH goal | 7.3 - 7.45 | 7.3 - 7.45 | 7.3 - 7.45 |
| Ventilator rate (breaths/min) | ≤ 35 | ≤ 35 | ≤ 35 |
| Oxygenation goal by PaO2 (mmHg) | 55 - 80 | 55 - 80 | 55 - 80 |
| Oxygenation goal by SpO2 (%) | 88 - 95 | 88 - 95 | 88 - 95 |
| Positive end-expiratory pressure | Set with FiO2-PEEP table | Set with FiO2-PEEP table | Set with FiO2-PEEP table |
| Inspiratory: Expiratory ratio | 1:1 - 1:3 | 1:1 - 1:3 | 1:1 - 1:3 |
| Extubation evaluation | MICU  SBT Readiness and Extubation Guidelines | MICU  SBT Readiness and Extubation Guidelines | MICU  SBT Readiness and Extubation Guidelines |

### 6. Guidelines for Spontaneous Breathing Trials and Ventilator Discontinuation in the Study ICU

**Part I – Spontaneous Breathing Trial Readiness Assessment**

Every morning or at least once daily, patients will undergo screening and trial in a Spontaneous Awakening Trial (SAT) performed by bedside nursing. If the patient passes the SAT, then the patient should be promptly evaluated in a Spontaneous Breathing Trial (SBT) Readiness Assessment as a safety screening:

If all of the following criteria have been present for more than 2 hours, then evaluate the patient for progression to Spontaneous Breathing Trial:

a)	FiO_2_ ≤ 0.5 & PEEP ≤ 8 cmH_2_O

b)	Values of both PEEP and FiO_2_ ≤ values from previous 12 hours.

c)	Not receiving neuromuscular blocking agents

d)	Patient is exhibiting adequate inspiratory efforts (if not, decrease the ventilator rate to 50% of baseline level for up to 5 minutes to detect inspiratory effort).

e)	Systolic arterial pressure > 90 mmHg without significant vasopressor support (< 10 mcg/min of norepinephrine or < 5 mcg/kg/min dopamine or dobutamine will not be considered a vasopressor).

**Part II – Spontaneous Breathing Trial (SBT)**

If criteria a-e above are met, then initiate a trial of at least 30 minutes of spontaneous breathing with FIO2 ≤ 0.5 using any of the following approaches:

1.	Pressure support ≤ 5cmH_2_O, PEEP ≤ 5cmH_2_O

2.	CPAP ≤ 5 cmH_2_O

3.	T-piece

4.	Tracheostomy mask

**Monitor for tolerance using the following:**

1.	SpO_2_ ≥ 88% and / or PaO_2_ ≥ 60 mmHg. FiO_2_ may be increased to 0.5 if necessary to maintain oxygenation in the target range.

2.	Mean spontaneous tidal volume ≥ 4 mL/kg PBW (if measured)

3.	Total Respiratory Rate ≤ 35 / min (< 5 min at RR > 35 may be tolerated, example post suctioning)

4.	pH ≥ 7.30 (if measured)

5.	No respiratory distress (defined as 2 or more of the following):

a.	Heart rate ≥ 120% of the 06:00 rate (≤ 5 min at > 120% may be tolerated)

b.	Marked use of accessory muscles

c.	Abdominal paradoxical breathing

d.	Diaphoresis

e.	Marked subjective dyspnea.

If any of the goals 1-5 are NOT met, revert to previous ventilator settings with Positive End-expiratory Pressure and FiO_2_ from the previous ventilator settings before the SBT was performed and reassess for the readiness for SBT the next morning.

The clinical team may decide to change mode of support during spontaneous breathing (PS = 5cmH_2_O, CPAP, tracheostomy mask, or T-piece) at any time.

**Part III – Decision to Remove Ventilatory Support**

For intubated patients, if tolerance criteria for spontaneous breathing trial (1-5 above) are

met for at least 30 minutes, continue with unassisted breathing, and extubate if clinically indicated under the direction of team physicians. However, the spontaneous breathing trial can continue for up to 120 minutes if tolerance remains in question (in fragile patients).

If any of criteria 1-5 are NOT met during spontaneous breathing (or 120 minutes has passed without clear tolerance), then the ventilator settings that were in use before the attempt to wean will be restored (Positive End-expiratory Pressure and FiO_2_ = previous settings) and the patient will be reassessed for an SBT the following day.

### 7. Guidelines for Assessment of Pain (CPOT score) in the Study ICU

| **Indicator** | **Assessment** | **Score** | **Description** |
| --- | --- | --- | --- |
| Facial expressions | Relaxed, neutral | 0 | No muscle tension observed |
|  | Tense | 1 | Presence of frowning, brow lowering, orbit tightening and levator contraction or any other change (e.g., opening eyes or tearing during nociceptive procedures) |
|  | Grimacing | 2 | All previous facial movements plus eyelid tightly closed (the patient may present with mouth open or biting the endotracheal tube) |
| Body movements | Absence of movements or normal position | 0 | Does not move at all (doesn’t necessarily mean absence of pain) or normal position (movements not aimed toward the pain site or not made for the purpose of protection) |
|  | Protection | 1 | Slow, cautious movements, touching or rubbing the pain site, seeking attention through movements |
|  | Restlessness/Agitation | 2 | Pulling tube, attempting to sit up, moving limbs/thrashing, not following commands, striking at staff, trying to climb out of bed |
| Compliance with the ventilator (intubated patients)  OR  Vocalizations (extubated patients) | Tolerating ventilator or movement | 0 | Alarms not activated, easy ventilation |
|  | Coughing but tolerating | 1 | Coughing, alarms may be activated but stop spontaneously |
|  | Fighting ventilator | 2 | Asynchrony: blocking ventilation, alarms frequently activated |
|  | Talking in normal tone or no sound | 0 | Talking in normal tone or no sound |
|  | Sighing, moaning | 1 | Sighing, moaning |
|  | Crying out, sobbing | 2 | Crying out, sobbing |
| Muscle tension  Evaluate by passive flexion and extension of the upper limbs when patient is at rest or evaluation when patient is being turned. | Relaxed | 0 | No resistance to passive movements |
|  | Tense, rigid | 1 | Resistance to passive movements |
|  | Very tense or rigid | 2 | Strong resistance to passive movements or incapacity to complete them |
| Total |  | /8 |  |

Adapted from: <https://www.icudelirium.org/medical-professionals/assess-prevent-and-manage-pain>

Gélinas, C. (2010). Nurses’ Evaluations of the Feasibility and the Clinical Utility of the Critical-Care Pain Observation Tool. Pain Management Nursing, 11(2), 115-125.

Arbour, C., & Gélinas, C. (2011). Ask the Experts. Setting Goals for Pain Management When Using a Behavioral Scale: Example With the Critical-Care Pain Observation Tool. Critical Care Nurse, 31, 66-68

### 8. Protocol for Assessment of Agitation (RASS Score) in the Study ICU

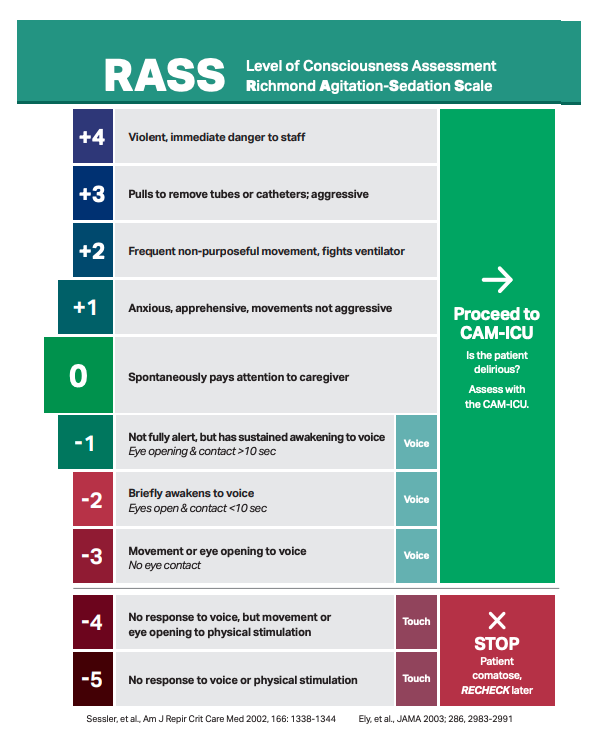

Reproduced with permission from: <https://www.icudelirium.org/medical-professionals/delirium/monitoring-delirium-in-the-icu>

### 9. Protocol for Delirium Assessment (CAM-ICU score) in the Study ICU

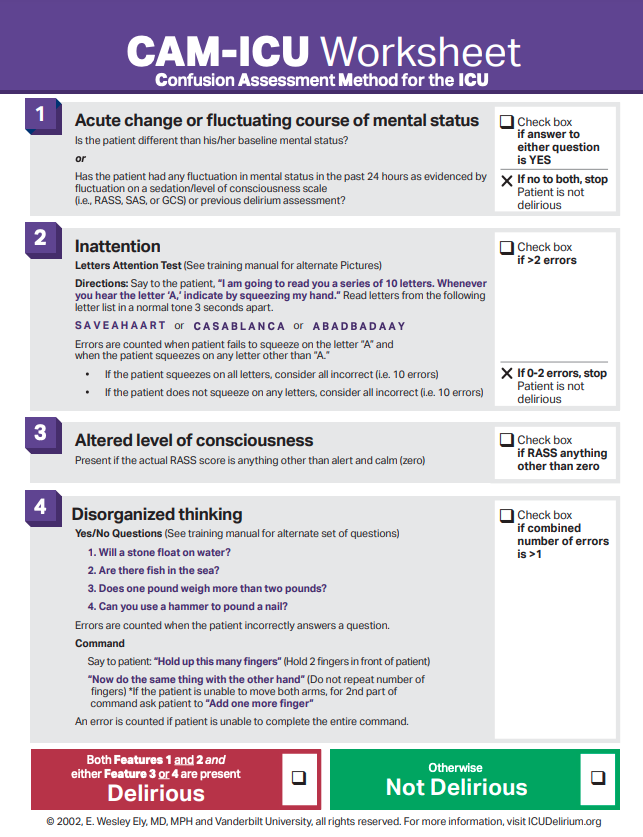

Reproduced with permission from: <https://www.icudelirium.org/medical-professionals/delirium/monitoring-delirium-in-the-icu>

### 10. Protocol for Management of Pain, Agitation, and Delirium in the Study ICU

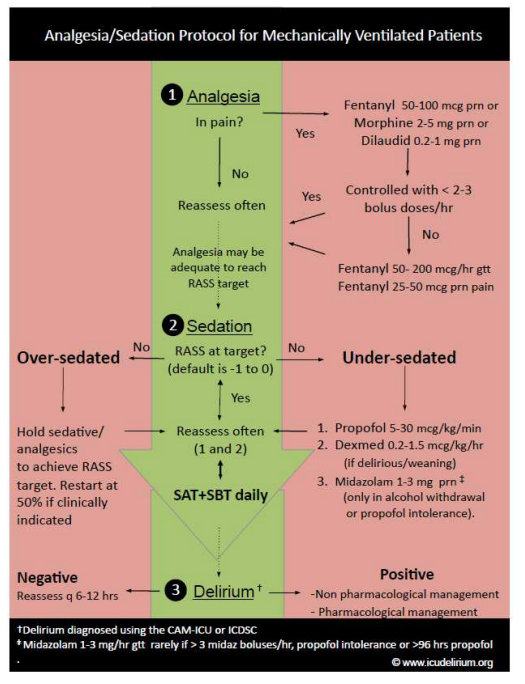

Reproduced with permission from: <https://www.icudelirium.org/medical-professionals/delirium/monitoring-delirium-in-the-icu>

### 11. Protocol for Early Mobility in the Study ICU

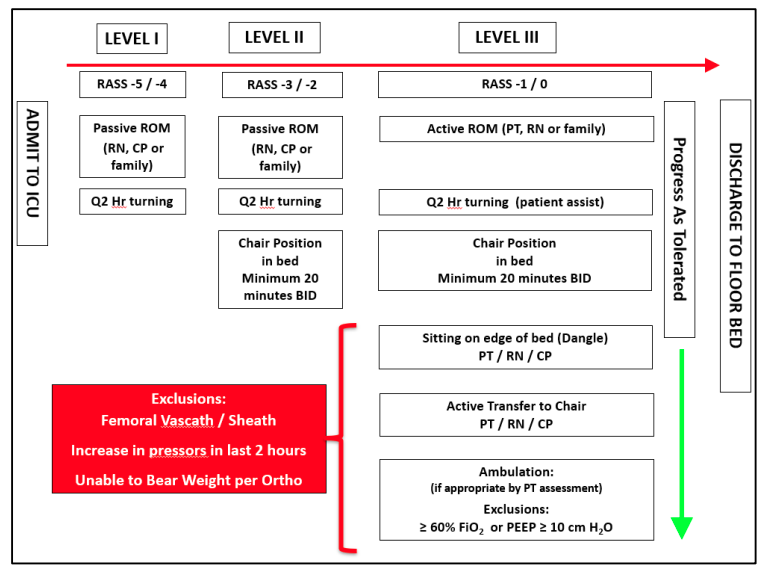

### 12. Treatment Decisions Determined by Treating Clinicians during the Study

All treatment decisions except choice of mode for continuous mandatory ventilation during invasive mechanical ventilation are made by treating clinicians, including: approach to supplemental oxygen therapy and non-invasive positive pressure ventilation prior to invasive mechanical ventilation, choice of oxygen saturation targets, use of spontaneous modes of ventilation such as pressure support, approach to positive end-expiratory pressure during invasive mechanical ventilation, prone positioning, or extracorporeal membrane oxygenation; administration of analgesia and sedation, neuromuscular blocking agents, inhaled epoprostenol, of vasopressors or inotropes, antimicrobial medications, diuretics, intravenous fluids, or blood products; measurement of lactate and arterial or venous blood gas.

### 13. Data Collected at Each Timepoint

Enrollment (Day 0)

1. Data collected by both manual and automated methods: age; sex; race; ethnicity; time from presentation to the study hospital to enrollment; time from ICU admission to enrollment; time from first receipt of invasive mechanical ventilation to enrollment; source of admission to the ICU;

2. Data collected only by manual method: baseline comorbidities; acute illnesses at enrollment; indications for invasive mechanical ventilation;

3. Data collected only by automated method: height, weight, Sequential Organ Failure Assessment (SOFA) score^11^; Elixhauser Comorbidity Index ^12^; Glasgow Coma Scale score^13^ ; vital signs (temperature, heart rate, systolic blood pressure, diastolic blood pressure, SpO_2_); mechanical ventilator settings (mode, set and exhaled tidal volume, set and actual respiratory rate, positive end-expiratory pressure, peak pressure, FiO_2_; serum laboratory values (white blood cell count, hemoglobin, platelet count, sodium, potassium, bicarbonate, creatinine, bilirubin, alanine aminotransferase, aspartate aminotransferase, lactate, arterial and venous measures of: pH, PaCO_2_, PaO_2_, SaO_2_).

On-Study (Days 0-28)

1. Data collected only by manual method:

a. Physician manual review blinded to study group assignment will determine whether each patient experienced acute respiratory distress syndrome (ARDS) by Berlin criteria^14^ pneumothorax or pneumomediastinum, use of prone positioning for hypoxemia.

b. For the full duration of the trial, each time treating clinicians elect to modify the assigned ventilator mode, study personnel will record the date and time of the modification, the mode to which the ventilator was changed, and the rationale for modifying the mode.

c. Each weekday, study personnel will record for each patient the results of the SAT Safety Screen, SAT, SBT Safety Screen, and SBT.

2. Data collected only by automated method: vital signs, ventilator settings, and serum laboratory values (as above); receipt of neuromuscular blockade; receipt of inhaled epoprostenol; number of arterial or venous blood gases; SOFA score; Richmond Agitation and Sedation Score^15^; Confusion Assessment Method for the ICU (CAM-ICU) score^16^.

Termination (Day 28)

Data collected by both manual and automated methods: death prior to hospital discharge; time from enrollment to death; duration of ICU admission; duration of hospital admission; duration of invasive mechanical ventilation; receipt of vasopressors; duration of vasopressor receipt; receipt of renal replacement therapy; duration of renal replacement therapy receipt; receipt of extracorporeal membrane oxygenation; duration of extracorporeal membrane oxygenation receipt.

### 14. Outcome Definitions:

Ventilator Free Days will be defined as the number of whole calendar days alive and free of invasive mechanical ventilation from the final receipt of invasive mechanical ventilation through day 28 after enrollment.^25,26^ Outcome ascertainment will cease at the time of hospital discharge or 28 days after enrollment, whichever occurs first. Receipt of invasive mechanical ventilation will be considered to end when patients undergo the final tracheal extubation or disconnection of the ventilator from the endotracheal tube or tracheostomy tube between enrollment (day 1) and day 28 day after enrollment. Patients who die prior to discharge on or before day 28 will receive 0 VFDs. Patients whose final receipt of invasive mechanical ventilation occurs on the day of enrollment and survive to day 28 will receive 27 VFDs. Patients who continue to receive invasive mechanical ventilation 28 days after enrollment will receive 0 VFDs. Patients who are discharged from the hospital prior to day 28 and are receiving invasive mechanical ventilation at the time of discharge will receive 0 VFDs. Patients who are removed from invasive mechanical ventilation and are discharged from the hospital without invasive mechanical ventilation prior to 28 days will be assumed to remain alive and free of invasive mechanical ventilation between hospital discharge and day 28. For patients who are removed from invasive mechanical ventilation, return to invasive mechanical ventilation, and are subsequently removed again from invasive mechanical ventilation prior to day 28, VFDs will be counted from the final receipt of invasive mechanical ventilation prior to day 28.

Median exhaled tidal volume (mL/kg PBW) on each study day: The median exhaled tidal volume on each study day will be defined as the median value of all recorded exhaled tidal volumes for the patient during each calendar day. Tidal volumes will be reported in mL/kg of Predicted Body Weight (PBW) by the following equations: PBW for Males = 50 + 2.3 * [height(inches)-60] and PBW for Females = 45.5 +2.3 * [height(inches) -60].

Delirium and Coma-Free Days through study day 28: Delirium and coma-free days are defined as the number of calendar days the patient is alive and free of delirium and coma from enrollment and day 28 after enrollment. Delirium is defined as a positive assessment of the CAM-ICU.^16^ Coma is defined as a RASS of -4 or -5.^15,17^ The number of days alive and free of delirium and coma is defined as the number of calendar days alive and free of both delirium and coma through day 28 using the “last off method.” The day of enrollment is considered to be day 1. Outcome ascertainment ceases at the time of hospital discharge or 28 days, whichever comes first. Delirium and coma-free days are considered to begin the day following the patient’s final recorded positive assessment of the CAM-ICU or RASS of -4 or -5 between enrollment and day 28. Patients who continue to have delirium or coma at day 28 after enrollment receive a value of 0. Patients who died on or before day 28 after enrollment receive a value of 0. Patients discharged from the hospital prior to day 28 after enrollment while still having delirium or coma receive a value of 0. Patients who were discharged from the hospital prior to day 28 after enrollment without delirium or coma are assumed to remain free of delirium and coma between hospital discharge and day 28 after enrollment. For patients who have a period without delirium and coma and subsequently developed or re-developed delirium or coma, days alive and free of delirium and coma are counted from the final day of delirium or coma through day 28 after enrollment.

ICU-free days through study day 28: Number of days alive and free of the ICU are defined as the number of calendar days in which the patient is alive between the patient’s final transfer or discharge from an ICU service and day 28 after enrollment. Patients who are never discharged from the intensive care unit receive a value of 0. Patients who die before day 28 receive a value of 0. Patients who return to an intensive care unit service and are subsequently discharged prior to day 28, ICU-free days are considered to begin the day following the date of final ICU discharge. All data are censored at hospital discharge or 28 days, whichever comes first.

Hospital-Free days to study day 28: Number of days alive and free of the hospital are defined as the number of calendar days between enrollment and 28 days after enrollment, in which the patient is alive and not admitted to the hospital. Patients who are never discharged from the hospital receive a value of 0. Patients who die before day 28 receive a value of 0. All data are censored at hospital discharge from the hospitalization in which they were enrolled in the trial or 28 days, whichever comes first.

### 15. Assessment of Compliance and the Assigned Study Mode

All ventilator modes collected during invasive mechanical ventilation are assessed for compliance with the assigned ventilator mode. A ventilator mode is considered compliant with the trial protocol if the following parameters are met:

1. The patient is receiving the study mode,
2. The mechanical ventilator is set on a mode for other than continuous mandatory ventilation, such as pressure support or synchronized intermittent mandatory ventilation,
3. The mode use occurs after treating clinicians have completed a Mode Modification Sheet.

### 16. Interim Analysis

The DSMB conducted a single, planned interim analysis for safety on March 22, 2023 to review data from patients enrolled after the ICU had been assigned to each of the three study modes, that is, after enrollment of patients for 3 months. According to criteria specified in the trial protocol, the stopping boundary for safety would have been met if the p-value for a difference between the groups in any of the three safety outcomes met the threshold of 0.016 or less. This priori threshold was determined by adjusting the p-value threshold of 0.05 for multiple comparisons by dividing 0.05 by the 3 outcomes. Given the design of the study is to demonstrate feasibility and pilot trial procedures rather than demonstrating efficacy, there was no stopping boundary for efficacy or futility.

The DSMB evaluated the incidence of the following safety outcomes across the study groups (volume control, pressure control, and adaptive pressure control): pneumomediastinum or pneumothorax during the course of mechanical ventilation, episodes of hypoxemia while receiving invasive mechanical ventilation: SpO_2_ < 85% for more than 5 minutes, and in-hospital mortality to study day 28.

After conducting the planned interim analysis, the DSMB recommended the study continue without modification. The DSMB reserves the right to stop the trial at any point, request additional data or interim analyses, or request modifications of the study protocol as required to protect safety.

### 17. Effect Modification Hypotheses

We have prespecified the following baseline variables as potential modifiers of the effect of study group on the primary outcome and hypothesized the direction of the effect modification for each:

1. Age (continuous variable). We hypothesize that age will not modify the effect of ventilator mode on the number of ventilator free days.
2. Duration of invasive mechanical ventilation prior to enrollment (0 minutes; 1 to 360 minutes; >360 minutes). We hypothesize that duration of invasive mechanical ventilation will not modify the effect of ventilator mode on the number of ventilator free days.
3. Pre-enrollment fraction of inspired oxygen (FiO_2_) (continuous variable). We hypothesize that the FiO2 received pre-enrollment will modify the effect of study mode on the outcome of ventilator free days with a greater increase in the number of ventilator free days in the volume control group among patients with a higher FiO2 compared patients with lower FiO2. This hypothesis is supported by the rationale that evidence-based practices for lung protective ventilation were established in prior trials that placed a priority on limiting tidal volumes, for which an effect was most pronounced among cohorts with severe hypoxemic respiratory failure.^4,5^
4. SOFA score at enrollment (continuous variable). We hypothesize that baseline SOFA score will not modify the effect of ventilator mode on the number of ventilator free days.
5. Shock receiving vasopressors (yes, no); We hypothesize that shock will not modify the effect of ventilator mode on the number of ventilator free days.
6. Indications for intubation (categories are not mutually exclusive)
   a. Hypoxemic respiratory failure (yes, no); We hypothesize that hypoxemic respiratory failure as a reason for intubation will modify the effect of ventilator mode on the primary outcome among the volume control group for the justification provided above with FiO2.
   b. Hypercarbic respiratory failure (yes, no); We hypothesize that hypercarbic respiratory failure will not modify the effect of ventilator mode on the number of ventilator free days.
   c. Altered mental status or airway protection (yes, no); We hypothesize that altered mental status or airway protection as a reason for intubation will modify the effect of study mode on the outcome of ventilator free days with a greater increase in the number of ventilator free days among the pressure control group for which improved patient-ventilator synchrony may increase patient comfort, reduce sedation use, and reduce the duration of mechanical ventilation.^28,29^
7. Chronic Obstructive Pulmonary Disease (yes, no); We hypothesize that chronic obstructive pulmonary disease will not modify the effect of ventilator mode on the number of ventilator free days.

### 18. Sensitivity Analyses of the Primary Outcome

We will repeat the primary analysis with adjustment for pre-specified baseline covariates of: age (continuous), sex (male, female), race and ethnicity (Hispanic, Non-Hispanic Black, Non-Hispanic White, Other), source of ICU admission (ED, hospital ward, another ICU in the study hospital, operating room, outside hospital), vasopressor receipt (yes, no), and acute diagnoses at enrollment (cardiac arrest, sepsis or septic shock, pneumonia, COPD exacerbation, asthma exacerbation), and severity of illness as assessed by SOFA score. To account for non-linear relationships, continuous variables will be analyzed using restricted cubic splines with between 3 and 5 knots. We will also repeat the analyses of the primary outcome and feasibility outcomes among all patients enrolled in the trial including those initiated on invasive mechanical ventilation in the study ICU during one of the pre-specified 3-day washout periods.

### 19. Handling of Missing Data

The primary outcome of VFDs is not anticipated to be missing for any patients. Missing data will not be imputed for the primary outcome or any exploratory outcomes. None of the covariates pre-specified for the adjusted analyses are anticipated to be missing for any patients. In additional adjusted analyses, any missing data for covariates will be imputed using multiple imputations.

### 20. Rationale for Waiver of Informed Consent

Volume control, pressure control, and adaptive pressure control are all common approaches to controlled mechanical ventilation for critically ill adults. All represent standard-of-care treatments in current clinical practice. Results from prior clinical trials do not demonstrate superiority of one approach over the other. Current clinical guidelines do not recommend any specific mode of mechanical ventilation.^18–21^ As a result, significant variation exists in the use of volume control, pressure control, and adaptive pressure control for patients in routine clinical care with ARDS^22^ and those without ARDS.^23^ In the study ICU, all three modes are used commonly in routine care. This trial enrolls only patients who would already receive continuous mandatory ventilation with one of these three modes as part of their routine clinical care. Whenever the treating clinicians feels that optimal care for the patient would involve a specific ventilator mode different from what is assigned by the study, the team can modify the ventilator mode for the patient and use the mode that they believe is optimal for the patient. Only patients for whom the treating clinicians feels that the ventilator mode assigned by the study is acceptable continue to receive the assigned ventilator mode. We requested and received a waiver of informed consent from the Vanderbilt University Medical Center Institutional Review Board (IRB 220446) because the study involves minimal risk and obtaining informed consent would be impracticable.

Participation in this study involves minimal risk because: First, the three interventions being compared are commonly used in routine clinical care in the study ICU. Second, all are interventions to which the patient could be exposed even if not participating in the study (all patients initiated on mechanical ventilation in the study ICU receive one of these modes of continuous mandatory ventilation). Third, no established differences in risk and benefits are known to exist between the studied approaches to mechanical ventilation based on the currently available data. Finally, the trial only determines the mode of ventilation when treating clinicians feel all three modes would be consistent with optimal care for the individual patient – otherwise treating clinicians select the mode of ventilation via a Mode Modification Sheet.

The MODE trial is designed as a cluster-randomized multiple-crossover trial to ensure that the mode selected at the time of initiation of mechanical ventilation is consistent with trial group assignment, preventing contamination between groups and capturing the period of mechanical ventilation in which ventilator settings have the strongest association with outcomes. Obtaining informed consent before initiation of mechanical ventilation in the study ICU would be impracticable because:

- The expected medical condition of patients at the time of initiation of invasive mechanical ventilation in the ICU is critical. All patients will be critically ill and receiving continuous intravenous sedation at the time of enrollment. Thus, all patients eligible for MODE will not have the capacity to provide informed consent. Further, data from prior trials in the same patient population and setting demonstrate that prior to intubation and initiation of mechanical ventilation, approximately 70% of patients eligible for the MODE trial will be experiencing encephalopathy (altered mental status) due to their illness. The anticipated median Glasgow coma scale score will be 11 (equivalent to moderate brain injury). Among the minority of patients whose level of consciousness is not impaired, 45-55% will be experiencing acute delirium. Further, family members or legally authorized representatives (LAR) are frequently unavailable when critically ill patients undergo intubation and initiation of invasive mechanical ventilation in the ED or ICU.
- The intervention is delivered by unit-level protocols. Mechanical ventilator settings and titrations are performed by respiratory therapists using unit-level protocols. In this cluster-randomized trial, the entire unit will be assigned to the same mode of mechanical ventilation for each 1-month block. Obtaining informed consent from every eligible patient in the ICU prior to emergency tracheal intubation and initiation of mechanical ventilation would be impracticable.

Because the study involves minimal risk, the study would not adversely affect the welfare or privacy rights of the participant, and obtaining informed consent would be impracticable, we will request a waiver of informed consent. Numerous previous randomized trials comparing two standards of care for interventions such as emergency intubation and methods of respiratory support during critical illness have been completed with waiver of informed consent.^10,24–31^

### 21. Plan for Communication of Protocol Changes

Any changes to the protocol (e.g., changes to eligibility criteria, outcomes, analyses) will require a new version of the full trial protocol which will be tracked with the date of the update and the version number of the trial protocol. A list summarizing the changes that are made with each protocol revision will be included at the end of each protocol. The updated protocol will be sent to the Vanderbilt University Medical Center IRB for approval prior to implementation of the protocol change. At the time of publication, the original trial protocol, and the final trial protocol, including the summary of changes made with each protocol change, will be included in the supplementary material for publication.

### 22. Patient Privacy and Data Handling

Patients will be followed after enrollment for 28 days or until hospital discharge, whichever occurs first. The minimum necessary data containing patient or provider identities will be collected. Data collected from the medical record and the treating clinician Mode Modification Sheets will be entered into the secure online database REDCap. Hard copies of the Mode Modification Sheet will be used for manual data entry into the REDCap database and stored in a locked room until after the completion of enrollment and data cleaning. All data will be maintained in the secure online database REDCap until the time of study publication. Only key study personnel will have access to the REDCap database. Once data are verified and the database is locked, all hard copies of Mode Modification Sheets will be destroyed. At the time of publication, a de-identified version of the database will be generated. At no time during this study, its analysis, or its publication, will patient identities be revealed in any manner.

### 24. Safety Monitoring and Adverse Events

Assuring patient safety is an essential component of this protocol. Mechanical ventilation with volume control, pressure control, and adaptive pressure control are all standard-of-care interventions that have been use in clinical practice for decades with an established safety profile. This protocol further ensures safety of its participants through:

a. Exclusion criteria designed to prevent enrollment of patients likely to experience adverse events from pressure control, volume control, or adaptive pressure control settings;
b. Permitting treating clinicians to change the ventilator mode at any time for any patient in whom they feel a ventilator mode different from the assigned mode is required for the safe treatment of the patient;
c. Systematic collection of outcomes relevant to the safety of ventilator mode selection;
d. Structured monitoring, assessment, recording, and reporting of adverse events.

**Adverse Event Definition–** An adverse event is defined as any untoward or unfavorable medical occurrence in a human subject temporally associated with the subject’s participation in the research, whether or not it is considered related to the subject’s participation in the research. Adverse events will be classified according to the following characteristics:

- **Seriousness** – An adverse event will be considered “serious” if it:
  - Results in death;
  - Is life-threatening (defined as placing the patient at immediate risk of death);
  - Results in inpatient hospitalization or prolongation of existing hospitalization;
  - Results in a persistent or significant disability or incapacity;
  - Results in a congenital anomaly or birth defect; or
  - Based upon appropriate medical judgment, may jeopardize the patient’s health and may require medical or surgical intervention to prevent one of the other outcomes listed in this definition.
- **Unexpectedness** – An adverse event will be considered “unexpected” if the nature, severity, or frequency is neither consistent with:
  - The known or foreseeable risk of adverse events associated with the procedures involved in the research that are described in the protocol-related documents, such as the IRB-approved research protocol; nor
  - The expected natural progression of any underlying disease, disorder, or condition of the subject experiencing the adverse event and the subject’s predisposing risk factor profile for the adverse event.
- **Relatedness** – The strength of the relationship of an adverse event to a study intervention or study procedure will be defined as follows:
  - Definitely Related: The adverse event follows (1) a reasonable, temporal sequence from a study procedure AND (2) cannot be explained by the known characteristics of the patient’s clinical state or other therapies AND (3) evaluation of the patient’s clinical state indicates to the investigator that the experience is definitely related to study procedures.
  - Probably or Possibly Related: The adverse event meets some but not all of the above criteria for “Definitely Related”.
  - Probably Not Related: The adverse event occurred while the patient was on the study but can reasonably be explained by the known characteristics of the patient’s clinical state or other therapies.
  - Definitely Not Related: The adverse event is definitely produced by the patient’s clinical state or by other modes of therapy administered to the patient.
  - Uncertain Relationship: The adverse event does not fit in any of the above categories.

**Monitoring for Adverse Events:** For patients whose mode of mechanical ventilation is controlled by the trial, investigators will evaluate for the occurrence of adverse events daily as part of the assessment of ventilator modes. Investigators will assess any adverse events that occur for whether the adverse event meets the criteria for recording and reporting outlined below.

**Recording and Reporting Adverse Events:** The following types of adverse events will be recorded and reported:

- Adverse events that are Serious and Definitely Related, Probably or Possibly Related, or of Uncertain Relationship.
- Adverse events that are Unexpected and Definitely Related, Probably or Possibly Related, or of Uncertain Relationship.

Adverse events that do not meet the above criteria will not be recorded or reported. Adverse events that the investigator assesses to meet the above criteria for recording and reporting will be entered into the adverse event electronic case report form in the trial database. The investigator will record a preliminary assessment of each characteristic for the adverse event, including seriousness, unexpectedness, and relatedness. For any adverse event that is **serious AND unexpected**, and definitely related, probably or possibly related, or of uncertain relationship, the investigator will report the adverse event to the principal investigator **within 24 hours** of the investigator becoming aware of the adverse event. For any other adverse event requiring recording and reporting, the investigator will report the adverse event to the principal investigator **within 72 hours** of the investigator becoming aware of the adverse event. The principal investigator will make the final determination regarding each characteristic for the adverse event, including seriousness, expectedness, and relatedness.

For adverse events that meet the above criteria for recording and reporting, the coordinating center will notify the DSMB, the IRB, and the sponsor in accordance with the following reporting plan:

| **Characteristics of the Adverse Event** | **Reporting Period** |
| --- | --- |
| Fatal or life-threatening (and therefore serious), unexpected, and definitely related, probably or possibility related, or of uncertain relationship. | Report to DSMB and IRB within 7 days after notification of the event. |
| Serious but non-fatal and non-life-threatening, unexpected, and definitely related, probably or possibly related, or of uncertain relationship. | Report to DSMB and IRB within 7 days of notification of the event. |
| All other adverse events meeting criteria for recording and reporting. | Report to DSMB and IRB as part of continuing review. |

The investigator will distribute the written summary of the DSMB’s periodic review of reported adverse events to the IRB in accordance with NIH guidelines (http://grants.nih.gov/grants/guide/notice-files/not99-107.html).

Clinical Outcomes that may be Exempt from Adverse Event Recording and Reporting: In this study of critically ill patients at high risk for death and other adverse outcomes due to their underlying critical illness, clinical outcomes, including death and organ dysfunction, will be systematically collected and analyzed for all patients. The primary and exploratory outcomes will be recorded and reported as clinical outcomes and not as adverse events unless treating clinicians or site investigators believe the event is Definitely Related or Probably or Possibly Related to the study intervention or study procedures. This approach – considering death and organ dysfunction as clinical outcomes rather than adverse events and systemically collecting these clinical outcomes for analysis – is common in ICU trials. This approach ensures comprehensive data on death and organ dysfunction for all patients, rather than relying on sporadic adverse event reporting to identify these important events. The following events are examples of study-specific clinical outcomes that would not be recorded and reported as adverse events unless treating clinicians or site investigators believe the event was Definitely Related or Probably or Possibly Related to the study intervention or study procedures:

- Death (all deaths occurring prior to hospital discharge or 28 days will be recorded);
- Organ dysfunction
  - Pulmonary – hypoxemia, hypercarbia, acute hypoxemic respiratory failure, pneumothorax;
  - Cardiac – hypotension, cardiac arrest, or shock with or without receipt of vasopressors;
  - Delirium or coma;
- Duration of mechanical ventilation;
- Duration of ICU admission;
- Duration of hospitalization.

Note: A study-specific clinical outcome may also qualify as an adverse event meeting criteria for recording and reporting. For example, a pneumothorax that the investigator considers Definitely Related to ventilator mode would be both recorded as a study-specific clinical outcome and recorded and reported as a Serious and Definitely Related adverse event.

**Unanticipated Problems involving Risks to Subjects or Others:** Investigators must also report to the principal investigator Unanticipated Problems Involving Risks to Subjects or Others (“Unanticipated Problems”), regardless of severity, associated with study procedures **within 24 hours** of the investigator becoming aware of the Unanticipated Problem. An Unanticipated Problem is defined as any incident, experience, or outcome that meets all of the following criteria:

- Unexpected (in terms of nature, severity, or frequency) given (a) the research procedures that are described in the protocol-related documents, such as the IRB-approved research protocol; and (b) the characteristics of the subject population being studied; AND
- Definitely Related or Probably or Possibly Related to participation in the research (as defined above in the section on characteristics of adverse events); AND
- Suggests that the research places subjects or others at a greater risk of harm (including physical, psychological, economic, or social harm) than was previously known or recognized.

Upon becoming aware of any event that may represent an Unanticipated Problem, the investigator will assess whether the event represents an Unanticipated Problem by applying the criteria described above. If the investigator determines that the event represents an Unanticipated Problem, the investigator will record the Unanticipated Problem in the Unanticipated Problem electronic case report form in the trial database. The investigators will obtain information about the Unanticipated Problem and report the Unanticipated Problem to the DSMB and the IRB within 15 days of becoming aware of the Unanticipated Problem.

### SUPPLEMENTAL REFERENCES

1.	Futier E, Constantin J-M, Paugam-Burtz C, et al. A Trial of Intraoperative Low-Tidal-Volume Ventilation in Abdominal Surgery. N Engl J Med 2013;369(5):428–37.

2.	Gajic O, Frutos-Vivar F, Esteban A, Hubmayr RD, Anzueto A. Ventilator settings as a risk factor for acute respiratory distress syndrome in mechanically ventilated patients. Intensive Care Med 2005;31(7):922–6.

3.	Fuller BM, Mohr NM, Drewry AM, Carpenter CR. Lower tidal volume at initiation of mechanical ventilation may reduce progression to acute respiratory distress syndrome: a systematic review. Crit Care 2013;17(1):R11.

4.	Needham DM, Yang T, Dinglas VD, et al. Timing of Low Tidal Volume Ventilation and Intensive Care Unit Mortality in Acute Respiratory Distress Syndrome. A Prospective Cohort Study. Am J Respir Crit Care Med 2015;191(2):177–85.

5.	Writing Group for the PReVENT Investigators, Simonis FD, Serpa Neto A, et al. Effect of a Low vs Intermediate Tidal Volume Strategy on Ventilator-Free Days in Intensive Care Unit Patients Without ARDS: A Randomized Clinical Trial. JAMA 2018;320(18):1872.

6.	The ICU-ROX Investigators and the Australian and New Zealand Intensive Care Society Clinical Trials Group. Conservative Oxygen Therapy during Mechanical Ventilation in the ICU. N Engl J Med 2020;382(11):989–98.

7.	Writing Committee and Steering Committee for the RELAx Collaborative Group, Algera AG, Pisani L, et al. Effect of a Lower vs Higher Positive End-Expiratory Pressure Strategy on Ventilator-Free Days in ICU Patients Without ARDS: A Randomized Clinical Trial. JAMA 2020;324(24):2509.

8.	Meade MO, Cook DJ, Guyatt GH, et al. Ventilation strategy using low tidal volumes, recruitment maneuvers, and high positive end-expiratory pressure for acute lung injury and acute respiratory distress syndrome: a randomized controlled trial. JAMA 2008;299(6):637–45.

9.	Kacmarek RM, Villar J, Sulemanji D, et al. Open Lung Approach for the Acute Respiratory Distress Syndrome: A Pilot, Randomized Controlled Trial*. Critical Care Medicine 2016;44(1):32–42.

10.	Semler MW, Casey JD, Lloyd BD, et al. Protocol and statistical analysis plan for the Pragmatic Investigation of optimaL Oxygen Targets (PILOT) clinical trial. BMJ Open 2021;11(10):e052013.

11.	Vincent J-L, Moreno R, Takala J, et al. The SOFA (Sepsis-related Organ Failure Assessment) score to describe organ dysfunction/failure: On behalf of the Working Group on Sepsis-Related Problems of the European Society of Intensive Care Medicine (see contributors to the project in the appendix). Intensive Care Med 1996;22(7):707–10.

12.	Elixhauser A, Steiner C, Harris DR, Coffey RM. Comorbidity Measures for Use with Administrative Data: Medical Care 1998;36(1):8–27.

13.	Teasdale G, Jennett B. ASSESSMENT OF COMA AND IMPAIRED CONSCIOUSNESS. The Lancet 1974;304(7872):81–4.

14.	Acute Respiratory Distress Syndrome: The Berlin Definition. JAMA [Internet] 2012 [cited 2023 Jul 5];307(23). Available from: http://jama.jamanetwork.com/article.aspx?doi=10.1001/jama.2012.5669

15.	Sessler CN, Gosnell MS, Grap MJ, et al. The Richmond Agitation–Sedation Scale: Validity and Reliability in Adult Intensive Care Unit Patients. Am J Respir Crit Care Med 2002;166(10):1338–44.

16.	Ely EW, Margolin R, Francis J, et al. Evaluation of delirium in critically ill patients: validation of the Confusion Assessment Method for the Intensive Care Unit (CAM-ICU). Crit Care Med 2001;29(7):1370–9.

17.	Ely EW, Truman B, Shintani A, et al. Monitoring sedation status over time in ICU patients: reliability and validity of the Richmond Agitation-Sedation Scale (RASS). JAMA 2003;289(22):2983–91.

18.	Barbas CSV, Ísola AM, Farias AM de C, et al. Brazilian recommendations of mechanical ventilation 2013. Part I. Rev Bras Ter Intensiva 2014;26(2):89–121.

19.	Fan E, Del Sorbo L, Goligher EC, et al. An Official American Thoracic Society/European Society of Intensive Care Medicine/Society of Critical Care Medicine Clinical Practice Guideline: Mechanical Ventilation in Adult Patients with Acute Respiratory Distress Syndrome. Am J Respir Crit Care Med 2017;195(9):1253–63.

20.	Griffiths MJD, McAuley DF, Perkins GD, et al. Guidelines on the management of acute respiratory distress syndrome. BMJ Open Resp Res 2019;6(1):e000420.

21.	Papazian L, Aubron C, Brochard L, et al. Formal guidelines: management of acute respiratory distress syndrome. Ann Intensive Care 2019;9(1):69.

22.	Qadir N, Bartz RR, Cooter ML, et al. Variation in Early Management Practices in Moderate-to-Severe ARDS in the United States. Chest 2021;S0012369221010783.

23.	Neto AS, Barbas CSV, Simonis FD, et al. Epidemiological characteristics, practice of ventilation, and clinical outcome in patients at risk of acute respiratory distress syndrome in intensive care units from 16 countries (PRoVENT): an international, multicentre, prospective study. The Lancet Respiratory Medicine 2016;4(11):882–93.

24.	Huang SS, Septimus E, Kleinman K, et al. Targeted versus Universal Decolonization to Prevent ICU Infection. N Engl J Med 2013;368(24):2255–65.

25.	Casey JD, Vaughan EM, Lloyd BD, et al. Protocolized Postextubation Respiratory Support to Prevent Reintubation: A Randomized Clinical Trial. Am J Respir Crit Care Med 2021;204(3):294–302.

26.	Kerlin MP, Small DS, Cooney E, et al. A Randomized Trial of Nighttime Physician Staffing in an Intensive Care Unit. N Engl J Med 2013;368(23):2201–9.

27.	Driver BE, Semler MW, Self WH, et al. Effect of Use of a Bougie vs Endotracheal Tube With Stylet on Successful Intubation on the First Attempt Among Critically Ill Patients Undergoing Tracheal Intubation: A Randomized Clinical Trial. JAMA [Internet] 2021 [cited 2021 Dec 28];Available from: https://jamanetwork.com/journals/jama/fullarticle/2787158

28.	Hillier TA, Pedula KL, Ogasawara KK, et al. A Pragmatic, Randomized Clinical Trial of Gestational Diabetes Screening. N Engl J Med 2021;384(10):895–904.

29.	Casey JD, Janz DR, Russell DW, et al. Bag-Mask Ventilation during Tracheal Intubation of Critically Ill Adults. N Engl J Med 2019;380(9):811–21.

30.	Silverberg MJ, Li N, Acquah SO, Kory PD. Comparison of Video Laryngoscopy Versus Direct Laryngoscopy During Urgent Endotracheal Intubation: A Randomized Controlled Trial*. Critical Care Medicine 2015;43(3):636–41.

31.	Janz DR, Casey JD, Semler MW, et al. Effect of a fluid bolus on cardiovascular collapse among critically ill adults undergoing tracheal intubation (PrePARE): a randomised controlled trial. The Lancet Respiratory Medicine 2019;7(12):1039–47.
